# An ergonomic assessment tool for evaluating the effect of back exoskeletons on injury risk

**DOI:** 10.1101/2021.07.22.21260715

**Authors:** Karl E. Zelik, Cameron A. Nurse, Mark C. Schall, Richard F. Sesek, Matthew C. Marino, Sean Gallagher

## Abstract

Low back disorders (LBDs) are a leading injury in the workplace. Back exoskeletons (exos) are wearable assist devices that complement traditional ergonomic controls and reduce LBD risks by alleviating musculoskeletal overexertion. However, there are currently no ergonomic assessment tools to evaluate risk for workers wearing back exos. Exo-LiFFT, an extension of the Lifting Fatigue Failure Tool, is introduced as a means to unify the etiology of LBDs with the biomechanical function of exos. We present multiple examples demonstrating how Exo-LiFFT can assess or predict the effect of exos on LBD risk without costly, time-consuming electromyography studies. For instance, using simulated and real-world material handling data we show an exo providing a 30 Nm lumbar moment is projected to reduce cumulative back damage by ∼70% and LBD risk by ∼20%. Exo-LiFFT provides a practical, efficient ergonomic assessment tool to assist safety professionals exploring back exos as part of a comprehensive occupational health program.

**HIGHLIGHTS:**

- Back exos are wearable assist devices that complement ergonomic controls for reducing low back disorder (LBD) risks
- However, no ergonomic assessment tools exist to evaluate LBD risks for workers wearing back exos
- We introduce Exo-LiFFT, an ergonomic assessment tool adapted from the Lifting Fatigue Failure Tool
- Exo-LiFFT is a practical tool that unifies the etiology of LBDs and biomechanical function of exos
- Exo-LiFFT can be used to assess or predict the effect of exos on LBD risk without EMG testing

## 1 INTRODUCTION

### 1.1 Low back disorders, ergonomics, & exoskeletons

Low back disorders (LBDs) are a leading cause of disability in the workplace, accounting for 38.5% of work-related musculoskeletal disorders reported in the U.S. (Bureau of Labor Statistics, 2016). Approximately one in four workers report having experienced low back pain in the previous three months (Luckhaupt et al., 2019; Yang et al., 2016). Worldwide, low back pain has been estimated to result in a loss of 818,000 disability-adjusted life years annually (Punnett et al., 2005). Overexertion due to lifting is a common source of work-related LBDs (Liberty Mutual, 2020).

Ergonomics practitioners have reduced work-related musculoskeletal disorder occurrences and costs (Burgess-Limerick, 2018; Goggins et al., 2008; Henshaw, 2002) by implementing interventions consistent with the hierarchy of controls (NIOSH/CDC, 2015). Ergonomic assessment tools have played an important role in identifying and mitigating risks. Nevertheless, medical costs, lost productivity, turnover, disability, and societal consequences due to LBDs remain high (Punnett et al., 2005; van der Wurf et al., 2021).

Exoskeletons (*exos*) are wearable assist devices that complement traditional ergonomic controls and could further reduce LBDs, and other work-related musculoskeletal disorders due to overexertion. Exos refer to a broad class of devices, ranging from passive (including quasi-passive mode-switching) to powered devices, and from rigid exoskeletons to soft exosuits (Fig. 1). Exos have been shown to reduce musculoskeletal forces (Bär et al., 2021; Howard et al., 2020; Kermavnar et al., 2020; Lamers and Zelik, 2021), a key risk factor for overexertion injuries in the workplace (Gallagher and Schall, 2017; Marras et al., 1993; Norman et al., 1998). Some early adopters have classified exos as personal protective equipment (PPE), while others consider them to be engineering controls, or tools. For instance, as of December 2020 Toyota North America was using exos as PPE, while Dow Chemical was using exos as engineering controls. These classifications reflect differences in how organizations choose to structure and manage their occupational health, safety, and ergonomics programs, and there are a number of legal and practical considerations that factor into why and when organizations define exos as PPE, or not (Peterson et al., 2020).

**Figure 1.**
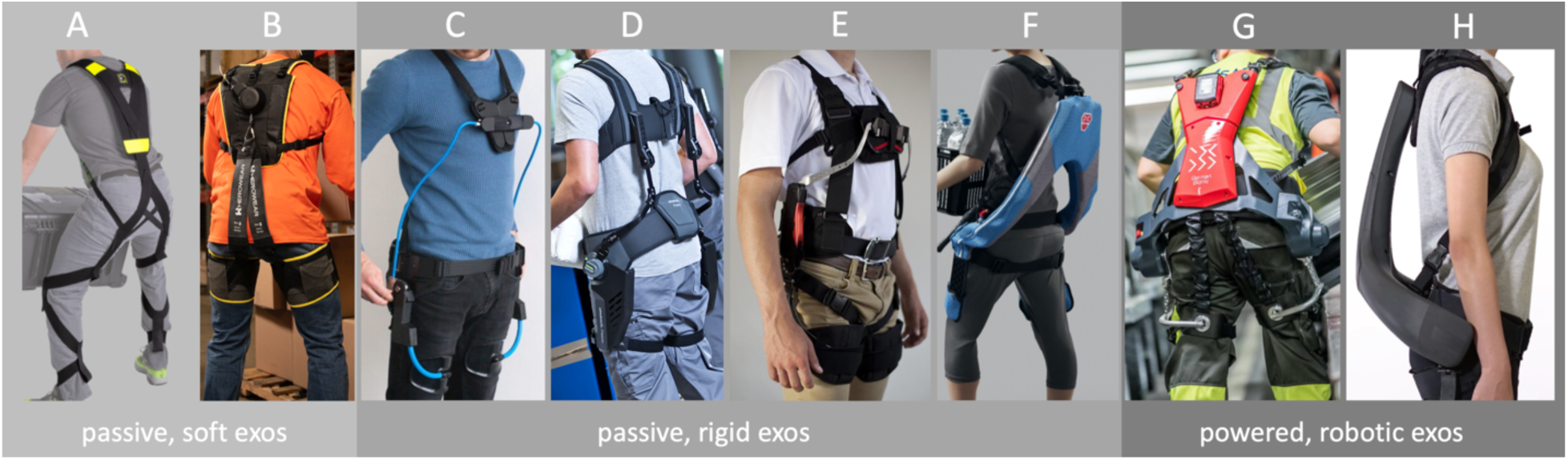
Examples of commercial back exos. Passive, soft exos (i.e., elastic exosuits made primarily from soft materials): A) Kinetic Edge Flex Lift, B) HeroWear Apex. Passive rigid exos (i.e., elastic exoskeletons made primarily from rigid components): C) Laevo, D) Ottobock Paexo Back, E) SuitX BackX, F) Innophys Muscle Suit Every. Powered robotic exos (i.e., using motors to assist): G) German Bionic Cray X, H) Atoun Model Y. Regardless of differences in componentry (rigid vs. soft) or actuation (passive vs. powered), each of these exos assists biomechanically by generating an extension moment (torque) about the lumbar spine during lifting, which reduces musculoskeletal loading on the user’s low back.

There is a growing interest in understanding and predicting how exos affect exposure to risk factors associated with work-related musculoskeletal disorders, such as LBDs from overexertion. Long-term injury data from large-scale, longitudinal epidemiological studies is sparse, and unlikely to become plentiful until exos are adopted at scale in natural environments, enabling evaluation of hundreds or thousands of workers over multiple years. Even then, data will be limited to exos already adopted in industry, leaving an unmet need for assessing the potential of new devices and for comparing jobs or tasks performed with and without exo assistance.

### 1.2 Prior success developing an exo-compatible ergonomic assessment tool for the shoulder

Researchers and safety professionals at Iowa State University and Lean Steps Consulting, in collaboration with safety managers at Toyota, adapted an established ergonomic assessment tool to investigate the effect of shoulder exos on injury risk (Butler and Gillette, 2019). This work was completed before longitudinal injury data were available to assess long-term exo effects. They used the Upper-Limb Localized Fatigue Threshold Limit Value (TLV) curves published by the American Conference of Governmental Industrial Hygienists (ACGIH, 2016), which provides recommended limits on the amount of time a worker should perform work at a given exertion level (e.g., due to postural demands) before experiencing excessive muscle fatigue. Reductions in upper limb muscle activity due to exo assistance were mapped onto the TLV curves to estimate expected reductions in muscle fatigue and injury risk (Butler and Gillette, 2019; Gillette and Stephenson, 2019).

These adapted TLV curves were used to provide an initial quantitative ergonomic assessment, indicating reduced shoulder muscle loads and risks for certain job tasks when workers wore an exo. This exo-compatible ergonomic assessment tool helped Toyota to deploy its resources efficiently, classify shoulder exos as PPE for certain job tasks, and initiate prolonged field tests. After a few years of exo use, Toyota has now reported lower medical costs and fewer injuries in job roles using shoulder exos (Barrero, 2019), providing long-term epidemiological evidence supporting the TLV-based risk predictions. This example highlights the benefits of academic-industry collaborations, and of adapting evidence-based, ergonomic assessment tools to be compatible with exos even before long-term injury data are available.

### 1.3 Ergonomic need & proposed solution for an exo-compatible risk assessment tool for the back

There are currently no ergonomic assessment tools designed or adapted to assess back injury risk when workers wear a back exo. Despite a large body of evidence and consistent findings that back exos reduce back muscle strain, muscle activity, and spine compression force (Bär et al., 2021; Howard et al., 2020; Kermavnar et al., 2020; Lamers and Zelik, 2021), there is currently no mapping of this musculoskeletal offloading effect into how much this may reduce damage to tissues – the underlying cause of overexertion injuries (Edwards, 2018; Gallagher and Schall, 2017) – or back injury risk. There is a pressing need for tools to estimate expected effects of exo assistance on LBD risk.

The Lifting Fatigue Failure Tool (LiFFT, Gallagher et al., 2017) could provide a unifying basis to assess or predict the effect of exo assistance on back injury risk during lifting-intensive material handling. LiFFT is based on fatigue failure processes, which underly how microdamage within materials (including biological tissues) accumulates during repeated loading cycles. Fatigue failure underlies both the etiology of overexertion injuries (Edwards, 2018; Gallagher and Heberger, 2013; Gallagher and Schall, 2017), and the biomechanical rationale for why/how exos are expected to reduce injury risks (Abdoli-Eramaki et al., 2007; Di Natali et al., 2021; Lamers et al., 2018), which is why this tool has the potential to unify ergonomic assessment and exo assistance. Here we describe how LiFFT can be adapted to assess injury risk when wearing back exos and share examples of various uses.

## 2 METHODS

### 2.1 LiFFT for ergonomic assessment of back injury risk and damage

LiFFT is a low back risk assessment tool that has been validated against two epidemiological databases, explaining 72–95% of the deviance in LBDs experienced by workers (Gallagher et al., 2017). LiFFT estimates *Cumulative Damage* (*D*) to the low back due to lifting, and associated back injury risk. For brevity, we use the term *LBD Risk* and equation variable *R* in this manuscript to refer specifically to the *probability of a job being a high-risk job*. This LBD Risk definition comes from the epidemiological data used to validate LiFFT (Marras et al., 1993; Zurada et al., 1997). Reductions in LBD Risk are correlated with reductions in actual low back injury incidence in the workplace, based on a multi-year prospective study (Marras et al., 2000).

LiFFT provides a simple, elegant and actionable tool using just two inputs: the number of lift repetitions (*n*), and the peak load moment (*M*_*pl,i*_) which is calculated for each (*i*^th^) lift as the weight of the object lifted multiplied by the peak horizontal distance from the lumbar spine to the load. The key LiFFT equations, adapted from (Gallagher et al., 2017), are as follows:

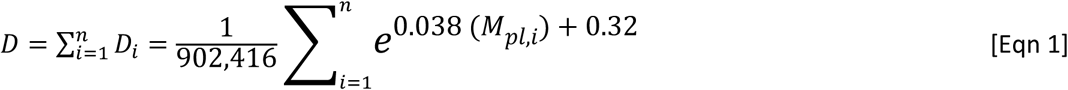

The scalar coefficient is derived from empirical fatigue testing on cadaveric spine specimens, and the exponent represents an empirically-derived relationship between peak load moment and compressive spine force as a function of ultimate strength. This equation was validated for peak load moments from 1.3-271 Nm (Gallagher et al., 2017), which corresponds with lifting roughly 0.5-80 kg. Logistic regressions and epidemiological databases with injury prevalence categorizations were then used to determine the relationship between Cumulative Damage (*D*) and LBD Risk (*R*):

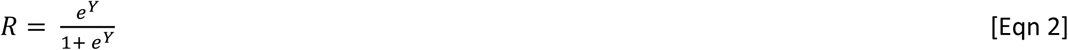

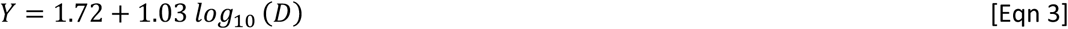

As with all ergonomic and biomechanical assessment tools, there are limitations to LiFFT. For instance, there are pros and cons to simplicity, such as using a discrete metric (peak load moment) to evaluate the risk induced by each lift, as opposed to using continuous data or more complex musculoskeletal load modeling or monitoring. The next several pages overview how LiFFT can be adapted to account for assistance from exos, followed by a detailed discussion in Section 3.5 of limitations, caveats, and other considerations when using this tool.

### 2.2 Exo-LiFFT for ergonomic assessment of back injury risk and damage while wearing exos

LiFFT equations can be modified to incorporate assistance from back exos. Back exos generate a moment (synonymous with the term *torque* herein) about the lumbar spine during lifting. Thus, it is possible to directly modify the peak load moment input to LiFFT by subtracting out the exo moment contribution from the peak load moment. In practice, this exo moment comes from mapping the lifting postures of workers (e.g., bend angles during lifting, as assessed by the safety professional) onto the mechanical assistance provided by an exo (e.g., back extension moment vs. joint angle curve provided by the exo manufacturer, or found from empirical testing of the device). The weight of the exo itself is also an important factor and its contributions to the lumbar moment should be accounted for in calculating the exo moment. For instance, if an exo has heavy componentry along the upper-body this would contribute a lumbar flexion moment which may partially counteract lumbar extension assistance from the exo’s actuator.

This modification of LiFFT is logical because the back moment provided by the exo reduces that moment borne by the biological back extensors (e.g., muscles, ligaments). This biological offloading is well supported by prior literature using electromyography (EMG), force-instrumentation, and musculoskeletal modeling (Bär et al., 2021; Howard et al., 2020; Kermavnar et al., 2020; Lamers and Zelik, 2021). For instance, Lamers et al. (2020) evaluated assistance benefits of a soft, passive-elastic back exo using two separate analyses – EMG, and a physics-based moment balance – and found the magnitude of back offloading estimated by each analysis to be similar. Likewise, a study on a rigid robotic back exo (Di Natali et al., 2021) found EMG and physics-based analyses yielded similar results in terms of reductions in back loading during lifting. In both studies, the physics-based analysis predicted slightly less back offloading than the EMG-based analysis, suggesting it may provide slightly more conservative estimates of exo benefits. Frost et al. (2009) demonstrated that as back exo assistance increased (from about 15 to 30 Nm) there was a proportional decrease in back muscle EMG, further corroborating the biomechanical function of back exos: moment demands on the biological back extensors are reduced by roughly the magnitude of moment provided by the exo, at least for mild to modest levels of assistance. By subtracting the exo back assistance moment from the peak load moment to estimate the ‘biological’ peak load moment, we can then input this updated peak load moment into LiFFT to compute Cumulative Damage and subsequently estimate LBD Risk while wearing an exo. The updated damage equation becomes:

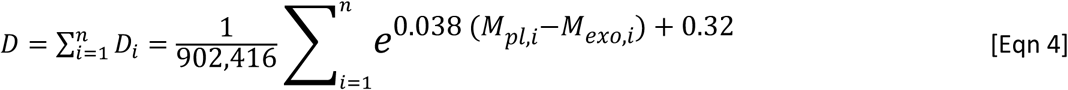

Where *M*_*exo,i*_ is the peak back extension moment provided by the exo on the *i*^th^ lift, and this equation is intended for 1.3 Nm ≤ (*M*_*pl,i*_ − *M*_*exo,i*_) ≤ 271 Nm based on prior LiFFT validation (Gallagher et al., 2017). The relationship between Cumulative Damage and LBD Risk remains unchanged from Eqns. 2-3.

We can quantify the change in Cumulative Damage and LBD Risk between exo (*D*_*exo*_, *R*_*exo*_) vs. no exo (*D*_*no_exo*_, *R*_*no_exo*_) conditions. Differences can be reported in non-normalized units (Eqns. 5-6), or it is often preferable to normalize differences to express them as a percentage of the no exo condition (Eqns. 7-8, e.g., for a safety professional seeking to understand how an exo affects injury risk relative to nominal work conditions without an exo):

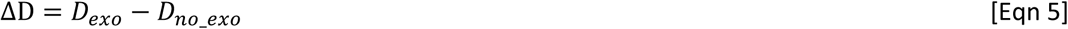

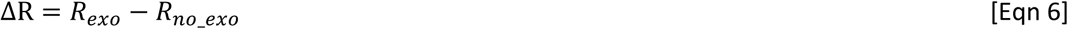

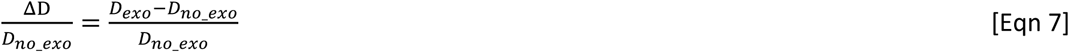

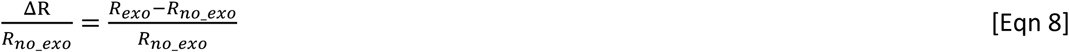

We refer to Eqns. 2-4, collectively, as Exo-LiFFT; a modification of the existing LiFFT that can be used to estimate effects of back exos. We report results from Eqns. 5-8 in terms of the percentage reduction magnitude due to exos; for instance, 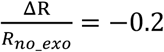 would be reported as a 20% reduction in LBD Risk. Next, we discuss three ways Exo-LiFFT can be used to predict or assess back exo effects on injury risk.

## 3 RESULTS & DISCUSSION

### 3.1 Simple prediction: Using Exo-LiFFT to estimate the effect of exo assistance on injury risk

It can be useful to quickly estimate the expected effect of an ergonomic intervention on injury risk. For instance, this could assist a safety professional doing a preliminary assessment of exos as a potential option to add to their operational toolbox. This use of Exo-LIFFT may involve identifying one or more jobs, then estimating risk reduction for a given exo, or comparing expected effects from multiple exos.

#### Example 1

Imagine a safety professional has already implemented good ergonomic practices within the hierarchy of controls, to the extent practical. However, back discomfort and injuries persist amongst workers. They decide to evaluate the potential benefits of a commercially-available back exo that provides 30 Nm of torque about the back during a typical lift. The safety professional already knows (from their previous ergonomic assessments, or from operational/organizational data) that their workers perform about 2,000 lifts per day, the average object lifted is 15 kg, and average distance from the spine to the object is about 50 cm. For this precursory analysis these values can be ballpark estimates, but note that the average peak exo moment (at or near the time of peak load moment) expected for a given job or task should be used, not simply the maximum moment an exo can generate. Given this simple information, the safety professional can now use Exo-LiFFT to estimate the Cumulative Damage and LBD Risk to the workers with vs. without the exo, to quantify expected effects of augmenting workers with exos. In this hypothetical example, Cumulative Damage and LBD Risk are expected to decrease by 67% and 20%, respectively, when using the exo (Fig. 2).

**Figure 2.**
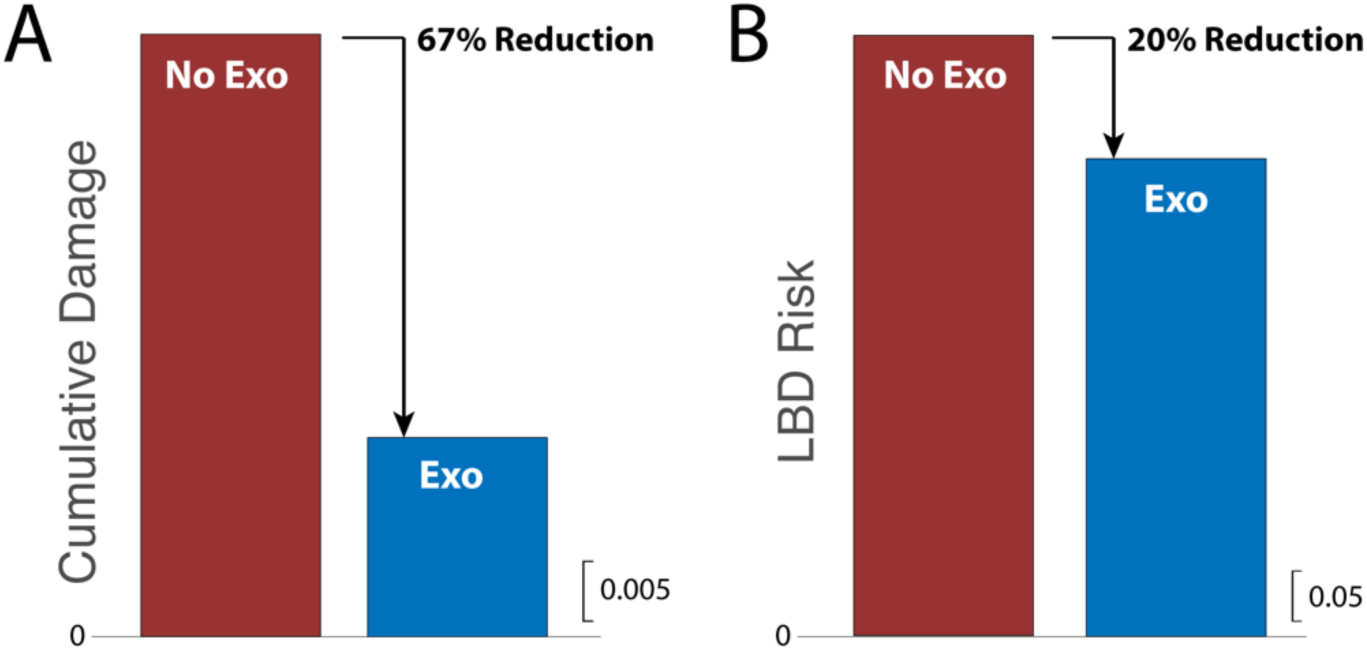
Exo-LiFFT simple prediction results from Example 1. A) Cumulative Damage is projected to decrease by 67%, and B) LBD Risk by 20% when using an exo providing a 30 Nm lumbar moment relative to not wearing the exo. Cumulative Damage and LBD Risk scales are shown (see Methods for formal definitions of each metric).

To expand upon Example 1, we performed similar analysis with updated parameters to assess how exo effects change with Light (5 kg), Moderate (12 kg; which corresponds with the Mixed category in Example 2), and Heavy (19 kg) objects, and with Mild (15 Nm), Moderate (25 Nm; corresponding with Mixed in Example 2), and Strong (35 Nm) exo assistance (Table 1).

**Table 1.** Simple prediction results. Reductions in LBD Risk are shown for different exo assistance and object weights. 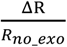 is reported as the primary summary metric, with ΔR in parentheses for reference. N/A indicates peak lumbar moment minus exo moment was outside the validated range of the model (see Methods for details).

|  | Light Objects<br>(5 kg) | Moderate Objects<br>(12 kg) | Heavy Objects<br>(19 kg) |
| --- | --- | --- | --- |
| **Strong Exo Assist**<br>**(35 Nm)** | N/A<br>(N/A) | 26.9%<br>(14.2) | 20.4%<br>(13.8%) |
| **Moderate Exo Assist**<br>**(25 Nm)** | 21.9%<br>(8.7%) | 19.7%<br>(10.6%) | 14.8%<br>(10.0%) |
| **Mild Exo Assist**<br>**(15 Nm)** | 14.5%<br>(5.8%) | 11.5%<br>(6.2%) | 8.4%<br>(5.7%) |

### 3.2 Simulated workdays: Using Exo-LiFFT to estimate a range of potential exo effects

Many jobs are highly variable such that the number of lifts and weights lifted differ day-to-day, or week-to-week. Construction work is one example, but there are also long cycle duration and highly variable jobs in agriculture, logistics, manufacturing, retail, military, and all other industries. For these jobs it may be beneficial to simulate a wide variety of scenarios, and then compute a range of exo effects. For example, one could simulate two extremes for a given job: the easiest expected lifting days (e.g., light weights with low repetition) vs. the hardest expected lifting days (e.g., heavy weights with high repetition). Alternatively, one could use Exo-LiFFT to explore thousands of randomly simulated workdays, as follows. First, one defines the range of relevant parameters for a given job, which may include the range (or distribution) of object weights and horizontal spine-to-object distances. Next, randomizing exo assistance (over a specified range) can be done to reflect that lifting a box from different heights/locations results in different bending postures and thus different amounts of exo assistance (e.g., based on an exo’s moment vs. joint angle behavior). Below we present one example of how to use Exo-LiFFT to assess a variety of simulated workdays.

#### Example 2

Imagine a safety professional seeks to understand the range of exo effects expected for a job whose lifting demands vary day-to-day. They estimate (or have data to indicate) that for a given job, a worker lifts 1,000-3,000 times each day, objects are typically 25-75 cm in front of the spine, and range from 2-22 kg in weight. If the objects are located at different heights within the workplace (e.g., ground vs. knee height), then assistance expected from a given exo will typically vary (e.g., with passive-elastic exos the assistive moment often increases with bend angle). Assume a given exo provides a peak moment between 10-40 Nm each lift. As a reminder, this exo assistance range would come from mapping the lifting postures of workers onto the mechanical assistance provided by an exo. Hundreds of simulated workdays can be created by randomly varying each of these parameters (i.e., summing over a series of randomized lifts to simulate each workday). Exo-LiFFT can then compute the expected reduction in Cumulative Damage and LBD Risk when wearing the exo each simulated workday. Results for 1,000 simulated workdays are depicted in Fig. 3. Regardless of variations in lifting intensity on a given workday (represented by standard deviation bars, Fig. 3), the percentage reduction in Cumulative Damage and LBD Risk were similar for each simulated workday: Cumulative Damage was projected to decrease by 58% ± 1%, and LBD Risk by 18% ± 1% when wearing vs. not wearing the exo.

**Figure 3.**
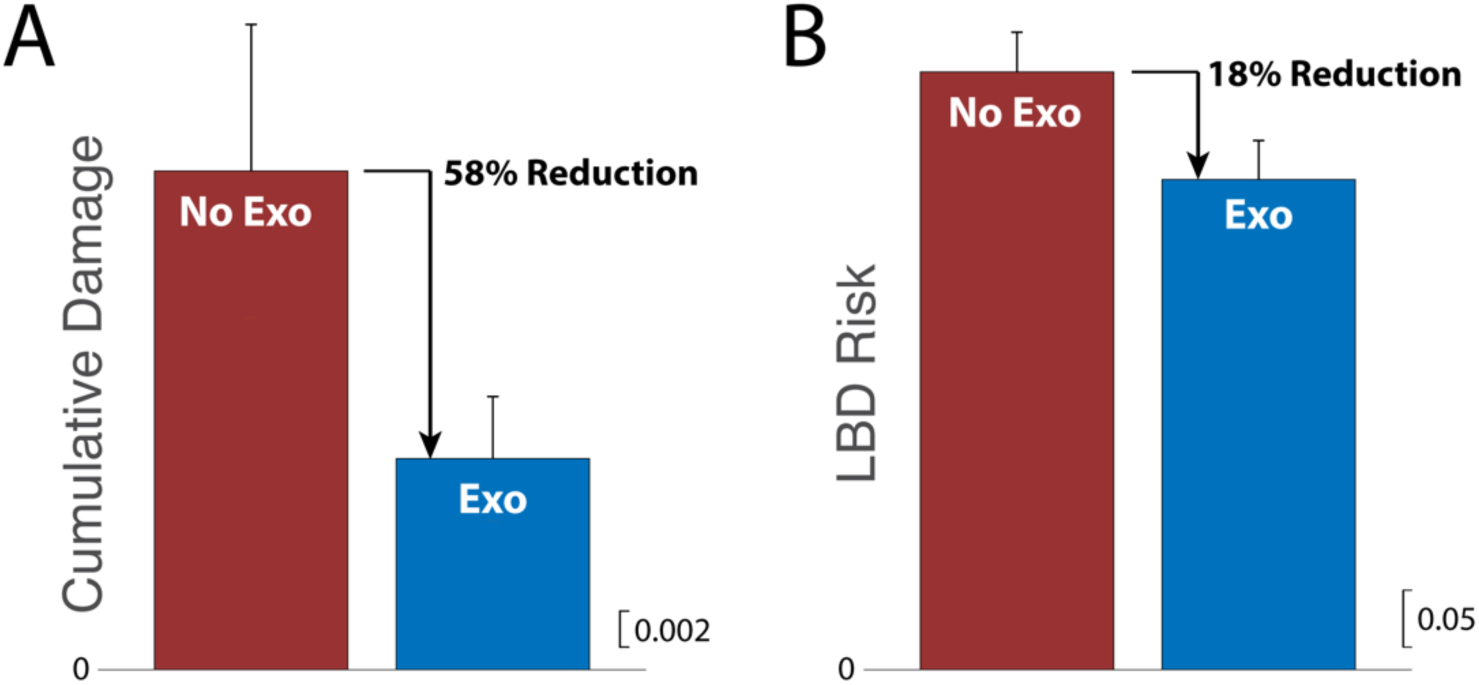
Exo-LiFFT results for 1,000 simulated workdays from Example 2. Means and standard deviations (across simulated workdays) are depicted. A) Cumulative Damage was estimated to decrease by 58%, and B) LBD Risk by 18%. Cumulative Damage and LBD Risk scales are shown (see Methods for formal definitions of each metric).

Next, we performed additional simulations with updated parameter ranges to assess how exo effects change with Light (2-8 kg), Heavy (16-22 kg), and Mixed (2-22 kg) objects, and with Mild (10-20 Nm), Strong (30-40 Nm), and Mixed (10-40 Nm) exo assistance (Table 2). The corresponding simple predictions using mean parameter values are shown in Table 1 (Moderate corresponds with Mixed).

**Table 2.** Simulated workday results. Reductions in LBD Risk are shown for different exo assistance and object weights. 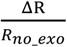 is reported as the primary summary metric, with ΔR in parentheses for reference. Each numerical result represents the mean and standard deviation from 1000 simulated workdays.

|  | **Light Objects**<br>**(2-8 kg)** | **Mixed Objects**<br>**(2-22 kg)** | **Heavy Objects**<br>**(16-22 kg)** |
| --- | --- | --- | --- |
| **Strong Exo Assist**<br>**(30-40 Nm)** | 24.0% ± 1.1%<br>(9.7% ± 0.4%) | 26.0% ± 1.4%<br>(13.6% ± 0.22%) | 26.2% ± 2.1%<br>(18.1% ± 0.7%) |
| **Mixed Exo Assist**<br>**(10-40 Nm)** | 18.8% ± 0.9%<br>(7.6% ± 0.3%) | 18.1% ± 1.1%<br>(9.5% ± 0.2%) | 15.8% ± 1.4%<br>(11.0% ± 0.5%) |
| **Mild Exo Assist**<br>**(10-20 Nm)** | 13.6% ± 0.7%<br>(5.5% ± 0.2%) | 11.7% ± 0.8%<br>(6.1% ± 0.1%) | 9.2% ± 0.9%<br>(6.3% ± 0.4%) |

Two key observations emerged from these simulated workdays. First, the reduction in LBD Risk is nearly constant for a given range of exo assistance and weights lifted (as evidenced by small standard deviations in Table 2), independent of the number of lift repetitions. This suggests that it is not necessary to precisely track every lift performed throughout a workday to obtain a reasonable estimate of exo effects on worker injury risk. Interestingly, even across the different object weights explored here, the percentage reduction in LBD Risk for a given range of exo assistance was also fairly similar. Second, LBD Risk magnitudes and trends from the simple prediction method (Table 1, range: 8.4%-26.9%, which used the mean parameter values only) match well those observed with the simulated workday analysis (Table 2, range: 9.2%-26.2%), reinforcing the utility and appropriateness of the quick, simple Cumulative Damage and LBD Risk predictions in Section 3.1.

Nevertheless, there may be scenarios where the range of parameter inputs is not known, or an exo is expected to substantially alter lifting behavior, or exos have already been adopted into regular use in the workplace. In these situations, it may be preferable to use real-world (non-simulated) movement and lifting data collected from workers on the job as inputs to Exo-LiFFT, as summarized below.

### 3.3 Ergonomic assessment: Using Exo-LiFFT to assess the effect of an exo worn in the workplace

Real-world data will capture any changes in behavior or lifting kinematics that may be user- or exo-specific. For instance, if an exo is too bulky it could force a worker to lift boxes further out from their body, which would increase the peak load moment of each lift relative to not wearing the exo. The drawback of collecting real-world data is that it is time-consuming and costly, and impractical to do for all users and jobs. Even optical and wearable sensor monitoring technologies still require professional oversight (Matijevich et al., 2021), and may also require customization or modification to accommodate exos. When data from actual workers is feasible to collect and process, such that key parameters can be extracted or estimated (e.g., lift repetitions, object weight, distance from spine to object, peak load moment, exo moment), then these data can be input into Exo-LiFFT instead of simulated data. If data are collected for workers during a period without an exo, and during a comparable period while wearing a given exo, then these can separately be input into Exo-LiFFT to quantify the differences. Alternatively, if data are only available from workers wearing an exo (e.g., if the exo is already adopted as part of their regular job), then one could compute Exo-LiFFT equations with vs. without exo assistance (analogous to comparisons in Sections 3.1-3.2).

#### Example 3

Real-world material handling data were analyzed and provided by HeroWear. Case pickers were recorded while wearing the Apex exosuit and performing their daily work tasks at the distribution center of a national retailer. Key parameters were extracted from video recordings of 210 lifts. Maximum bending (trunk-to-thigh) angle was measured using a goniometer (61° ± 23°, mean ± standard deviation, where 0 degrees represents upright standing, and this angle increases as a person bends forward). Peak horizontal distance from the L5-S1 spine to the object was measured (56 ± 13 cm) using a visual look-up table of squatting and stooping postures with associated horizontal distance measurements. Object weight (18 ± 5 kg) was found by looking up products on the retailer’s website. When products were not visible or bending angles were not directly measurable due to the camera angle, then values were estimated by a Certified Professional Ergonomist and/or Physical Therapist familiar with material handling in logistics and retail. Maximum bending angle was used with exo moment-angle curves from the exo manufacturer to compute the peak exo assistance for each lift. Peak exo moment was computed for two different elastic stiffness levels (23 ± 5 Nm for the Strong assistance bands, 30 ± 8 Nm for the Extra Strong assistance bands provided by the manufacturer). Cumulative Damage and LBD Risk were then computed, assuming these representative lifts continued for a worker’s typical shift (8 hours at a rate of 175 lifts per hour). When wearing the exo Cumulative Damage decreased by 58% and 73%, and LBD Risk decreased by 14% and 22%, respectively, for the two exo assistance levels (Fig. 4). The results from the real-world exo assessment match closely the estimates from the simple prediction method (Fig. 2, Table 1) and the workday simulations (Table 2). For instance, results from the 30 Nm exo in this real-world assessment (73% reduction in Cumulative Damage, and 22% reduction in LBD Risk, Fig. 4) were similar to those from the 30 Nm hypothetical exo in the simple prediction example (68% and 20% reductions, respectively, Fig. 2).

**Figure 4.**
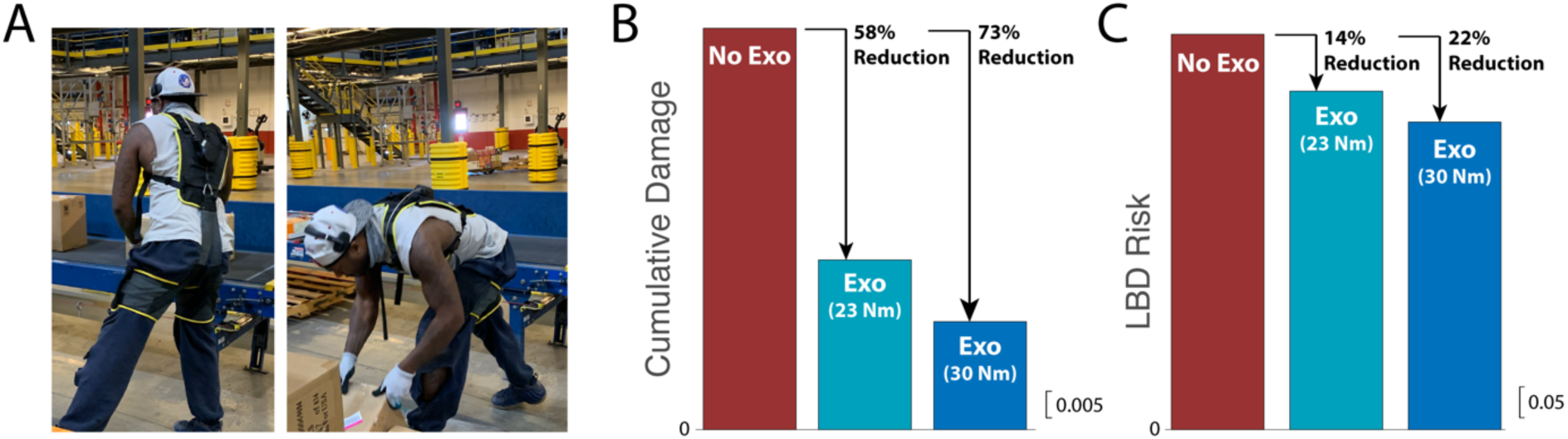
Real-world case study results on logistics case pickers wearing an exo. Example 3 results are shown for A) the HeroWear Apex exosuit, configured with Strong and Extra Strong assistance bands. B) Cumulative Damage decreased by 58% and 73%, and C) LBD Risk decreased by 14% and 22%, respectively, relative to not wearing an exo. Cumulative Damage and LBD Risk scales are shown (see Methods for formal definitions of each metric).

Reductions in LBD Risk and lumbar moment due to the exo in Example 3 are similar to those previously estimated for ergonomic lift tables. Lift tables were found to reduce lumbar moments by 35 Nm and LBD Risk by 25% (Marras et al., 2000), which is similar to the 30 Nm and 22% LBD Risk reductions from the exo in Example 3 (and also similar to the hypothetical exo in Example 1). In the Marras et al. (2000) prospective workplace injury study, lift tables reduced LBD incidence rates by about 7.5 LBDs per 100 full-time employees (in jobs averaging 12 LBDs per 100 employees per year); signifying about a 60% reduction in actual LBDs at work. Regression models based on a larger ergonomics and epidemiological data set (Marras et al., 2000) predict an injury reduction of about 20-50% (2.5-6 LBDs per 100 employees) when LBD Risk is reduced by the magnitudes in exo Examples 1 and 3 (with 30 Nm of exo assistance). This range of injury reduction overlaps with the Exo-LiFFT prediction. Thus, these epidemiological data provide additional support that the LBD Risk metric from Exo-LiFFT is a reasonable, and potentially somewhat conservative, prediction of exo effects on workplace back injuries. Collectively, these findings suggest that exos such as those exemplified here may have potential to reduce back injury incidence by roughly 20-60% in material handling (e.g., a reduction of 2.5-7.5 LBDs per 100 employees doing jobs averaging 12 LBDs per year). This projection should be evaluated in future large-scale, longitudinal, prospective studies on exos.

### 3.4 The need & benefit of keeping ergonomic assessment tools as simple as practical

Historically, back exo research, including our own (Lamers et al., 2020, 2018), has relied heavily on EMG assessment. A likely question from readers may be: why analyze lumbar moments rather than changes in back muscle EMG? The short answer is because these two methods have generally yielded similar results and conclusions for back exos (see Section 2.2); however, in our experience, exo EMG studies are much more complicated, time-consuming, invasive, and expensive to complete. We expect that using lumbar moments to assess back exo effects will be simpler, quicker, less costly, and more easily generalizable across exos than EMG, while generally providing similar insights on injury risk. It is preferable to keep ergonomic assessments as simple as practical to maximize accessibility to safety professionals and benefits to workers and organizations.

EMG is a great tool for research but is difficult to use as the basis for accessible, scalable, practical ergonomic assessment, particularly for the back. To briefly summarize key challenges: (i) EMG requires considerable time and expertise to collect and process (Besomi et al., 2020), and as such this level of analysis is not accessible or affordable for most safety professionals; (ii) there are an abundance of back extensor muscles, many of which are not measurable with surface EMG, meaning EMG studies only capture a subset of back muscle behaviors; (iii) there is substantial inter-subject variability with respect to which back muscles individuals offload and in what proportion when exo assistance is provided (Lamers et al., 2020), meaning it is unclear which subset of muscles to measure or how to make equitable comparisons between different individuals or exos; (iv) EMG only captures changes in muscle activity, and fails to capture reductions in spinal ligament loading (e.g., during stooped bending); (v) the relationship between EMG and force is non-linear, and dependent on other factors like muscle length, contraction velocity, electrode placement, and body posture (Ranavolo et al., 2020; Trinler et al., 2018), which can confound exo vs. no exo comparisons if kinematics are not tracked or controlled (Koopman et al., 2020a; Lamers et al., 2020); (vi) there can be safety concerns with normalization procedures that require maximum contractions (Cholewicki et al., 2011); and (vii) there is within- and between-participant variability when measuring EMG (e.g., due to motion artifacts, tissue conductivity) even without exos (Cholewicki et al., 2011), which can introduce reliability or interpretation issues, and these must be overcome with rigorous experimental design and data processing (Besomi et al., 2020).

EMG seems best suited for experienced researchers or safety professionals that seek to deeply study the effect of a given exo on a specific subset of muscles or users, during specific tasks or postures, or might potentially be used for other applications such as exo control (Coker et al., 2021). But we caution against over-reliance on EMG for assessing ergonomic effects of back exos due to the time, cost, and complexity burden it places on safety professionals and companies. To date, much of the back exo EMG research boils down to a confirmation that the lumbar moment borne by the musculoskeletal system is reduced by the magnitude of moment provided by the exo (e.g., Di Natali et al., 2021; Lamers et al., 2020), and that for passive exos inserting a stiffer spring generates more force and thus a larger exo moment (e.g., Frost et al., 2009).

We recommend the use of physics-based (lumbar moment) analysis for assessing back exos. This is particularly prudent for devices or assistance levels where EMG studies have already been conducted that confirm consistency with moment-based estimates. Lumbar moment analysis is well justified to use for evaluating exos that do not result in substantial co-contraction of the abdominal muscles or major changes to lifting kinematics; which seems to be the case for most current commercial exos, as detailed in the following paragraphs. However, for completely new classes of back exos without prior experimental studies, or unique new tasks that are dissimilar to typical lifting/bending activities, or much higher levels of assistance than provided in current commercial exos, or exos that introduce movement restrictions, then a more cautious and costly approach may be warranted (e.g., involving EMG, kinematics, and kinetics evaluation).

Most studies on back exos have not observed abdominal muscle activity to increase during lifting or bending (Kermavnar et al., 2020). For instance: Abdoli-E et al. (2006), Lamers et al. (2020), and Goršič et al. (2021) all found no substantial changes to abdominal muscle activity with vs. without a soft, passive-elastic back exo, and that abdominal EMG did not exceed 5% maximum voluntary contraction with or without the exo. While wearing a similar exo, Frost et al. found no differences in rectus abdominis EMG regardless of lifting technique, and no differences in external oblique EMG during squat or freestyle lifts, but did observe an increase in external oblique EMG during stoop lifting with the exo. When wearing a back exo comprised of carbon fiber beams, Baltrusch et al. (2020) found no change in abdominal muscle EMG, whereas Alemi et al. (2019) observed a slight increase in external oblique EMG during lifting. While wearing a rigid back exo during lifting, Koopman et al. (2020a) and Madinei et al. (2020) both found no difference in abdominal EMG, while Luger et al. observed a slight decrease in abdominal EMG. This list is not comprehensive, but is representative of current evidence (e.g., see systematic review by Kermavnar et al., 2020).

The degree to which lifting kinematics change or do not change depends on the type of exo and task (Kermavnar et al., 2020), but most evidence suggests that minor or no changes in posture occur when using current back exos (Madinei et al., 2020). For instance: Goršič et al. (2021) found no differences in 23 of 24 kinematic comparisons across a series of six lifting/lowering tasks while wearing a soft, passive-elastic back exo. The only difference was a slight decrease (of 5° or 8%) in trunk flexion/extension range-of-motion while wearing the exo during one asymmetric lifting task. For a similar type of exo, Abdoli-Eramiki et al. (2006) reported minor or no kinematics differences in lumbar flexion, pelvic flexion, or load acceleration, but did observe a slight reduction in trunk acceleration when wearing the exo for a subset of squat lifting tasks. In a follow-on study, Sadler et al. (2011) found that this exo reduced lumbar flexion and increased hip flexion, which the authors interpreted as evidence that the exo encouraged safe lifting practices without adversely affecting lifting technique. Baltrusch et al. (2020) tested a back exo using a carbon fiber beam and did not find that it significantly altered lifting kinematics. Kim et al. (2020) and Madinei et al. (2020) evaluated two rigid back exos and also found no substantial effects on working postures, or on triaxial trunk or lumbar ranges of motion. Lumbar flexion angles with these exos typically changed by less than 8° relative to not wearing an exo. They did observe a slight reduction (7%) in trunk velocity during lifting when wearing one of the two back exos tested, which was qualitatively consistent with Koopman et al. (2020a) who observed an 18% reduction in peak trunk velocity while wearing the same exo. Almosnino et al. (2021) also found that the time to reach peak trunk flexion was slightly delayed when wearing a rigid back exo, but concluded that it did not alter inter-segmental coordination of the pelvis-trunk during lifting. Koopman et al. (2020b) observed differences in lifting kinematics but concluded that these did not modify the biomechanical effect of the exo on low back loading. Similarly, Luger et al. (2021) found that a rigid exo only led to small changes in knee and hip flexion on the order of 2-5°, and did not lead to any impairments in spinal posture, which the authors concluded was in line with previous back exo studies. This summary is not meant to be comprehensive but simply to provide a snapshot of the current state of evidence suggesting that most current back exos have negligible or minor effects on lifting kinematics.

### 3.5 Limitations, caveats, & other considerations

Exo-LiFFT fills a contemporary ergonomics need by providing an estimate of exo effects based on the etiology of overexertion injuries, in conjunction with existing exo and epidemiological evidence. As long-term injury data emerge then Exo-LiFFT could be further refined, and predictions could be calibrated, validated or improved using larger datasets that contain epidemiological data (e.g., by determining if the constants from LiFFT remain similar or if they should be updated when wearing exos or for specific jobs/industries). In the meantime, the key to using Exo-LiFFT is being transparent and realistic in defining input parameters/ranges, then stating assumptions and confirming they are reasonable or well-supported for a given exo and use case. Exo-LiFFT can be used to assess exo effects on back injury risk, which complements evaluation of other factors (e.g., cost, weight, comfort, fit, freedom-of-movement, complexity, implementation, maintenance) that affect user experience and organizational adoption.

Exo-LiFFT is intended to assess lifting-intensive jobs that result in mechanical fatigue due to repetitive tissue loading. Exo-LiFFT in its current form is not intended for assessing risks due to prolonged static postures, such as sustained bending. In the future this tool might be extended to incorporate mechanical creep damage due to sustained lumbar forces; however, this would require additional data inputs, and may provide limited new insight for material handling because mechanical fatigue is expected to be the dominant mechanism of accumulated tissue damage, except at low levels of tissue loading (Gallagher and Huangfu, 2019). Nonetheless, Exo-LiFFT may underestimate the beneficial effect of back exos for workers who engage in sustained bending postures. Alternatively, Exo-LiFFT may overestimate back injury risk reduction if a given exo interferes with movement, the surrounding environment, or the ability of workers to complete job tasks normally (e.g., due to changes in lift distances, kinematics, task duration, or movement restrictions). A simple way to assess whether or not movement is restricted is to have workers wear a given exo during job tasks and then self-report if they feel the device interferes with movement, affects effort required to perform different tasks, or inhibits their ability to move freely (Baltrusch et al., 2018; Goršič et al., 2021; Yandell et al., 2021).

Exo-LiFFT is not intended to evaluate changes in risk or damage to other body parts. For a given exo, injury risk may increase or decrease at other body parts. However, we caution against assuming that an exo that reduces forces on the low back must therefore increase (or transfer) forces to other biological joints. This is a common misconception that often stems from misunderstanding the physics of how exos provide assistance, which is by using leverage to provide joint torque with a lower force magnitude than would be required by muscles or ligaments (Zelik, 2020). Neither muscle force nor joint contact force (Vigotsky et al., 2019) inside the body is a conserved quantity. There is no fixed magnitude of force that must be distributed over muscles or joints. The forces experienced by muscles, discs, and other tissues are a function of, amongst other factors, the mechanical leverage used by muscles, ligaments, or exos to generate torque (Abdoli-Eramaki et al., 2007; Lamers and Zelik, 2021).

There are various other ways to add complexity to the Exo-LiFFT model in terms of simulating different movements or scenarios, computing effects on different body parts, or calculating biomechanical moments using different models or assumptions (e.g., Merryweather et al., 2008). In the future, Exo-LiFFT might, for instance, be refined to incorporate factors such as trunk flexion, asymmetry or individual characteristics (e.g., height, weight, age, gender), which are not part of the current tool/model. However, it should be recognized that the simple underlying tool (LiFFT) has shown dose-response relationships to multiple low back outcomes in two separate epidemiological databases in which dynamic lifting was performed (Gallagher et al., 2017). In general, changes to the tool would require additional information or model validation before implementing. From the perspective of achieving a versatile, scalable, accessible ergonomic tool adding more complexity is not highly desirable.

This discussion of limitations and caveats is not comprehensive. Researchers and safety professionals should use good judgement and be attuned to circumstances that may preclude the use of Exo-LiFFT. Simple examples may include exo users that are reporting they are uncomfortable, or fighting against a particular exo to move, or unable to complete their job tasks due to exo weight, bulk, control, complexity, movement restriction or interference.

## 4 CONCLUSION

Exo-LiFFT (an adaptation of LiFFT) provides a practical, efficient, and informative ergonomic assessment tool to assist safety professionals exploring back exos as part of a comprehensive occupational health program. Exo-LiFFT unifies the etiology of LBDs due to overexertion with the biomechanical function of back exos, enabling an evidence-based way to convert exo assistance (lumbar moments) into expected effects on Cumulative Damage and LBD Risk. Using lumbar moments provides a practical alternative to costly and time-consuming EMG studies. Quick, simple predictions using Exo-LiFFT were similar to those obtained from simulated workdays and from real-world exo assessments, highlighting the utility of this tool and indicating that precise tracking of every lift a worker does during a workday is not necessary to estimate the effect of exos on injury risk during material handling or other repetitive lifting jobs.

## Data Availability

Data are available from the corresponding author, K.E.Z., upon reasonable request.

## ACKNOWLEDGEMENTS

The study was partially supported by the NIH (R01EB028105), and by the Deep South Center for Occupational Health and Safety, a NIOSH Education and Research Center (5 T42 OH 008436-16). The contents are solely the responsibility of the authors and do not represent official views of the Deep South Center, NIOSH, or NIH. These funding sources were not involved in study design, in the collection, analysis or interpretation of data, or in the decision to submit this article for publication. We gratefully acknowledge helpful feedback from Jason Gillette and Terry Butler, and thank Gavin Lee for assistance with data analysis.

### APPENDIX

#### A.1 Alternative approaches to assessing or communicating risk reduction from wearing exos

Before adapting LiFFT to accommodate exo assistance we considered a number of other ergonomic assessment tools or approaches. Here we briefly summarize our thoughts on these alternatives.

One alternative way to express exo assistance is in terms of “equivalent weight” or “equivalent effort” metrics (Di Natali et al., 2021; Lamers et al., 2020). For instance, instead of reporting Cumulative Damage or LBD Risk reduction, Example 1 could be summarized by stating that the 30 Nm back exo makes lifting a 15 kg box feel like lifting a 6.4 kg box from the standpoint of loading experienced by the back. Or alternatively, the exo makes lifting a 15 kg box 3,000 times feel like only lifting it 1,000 times, again from the perspective of back loading. This way of reporting exo effects is intuitive, and in our experience can be an effective and useful way to communicate expectations to broad audiences, which is consistent with (i.e., has the same physics-based rationale as) the Exo-LiFFT approach we propose here. However, this “equivalent weight” approach stops short of providing quantification of effects on injury risk, and does not map exo effects into any existing ergonomic guidelines or frameworks. As such, it is limited in its ability to help safety professionals formally assess, document, or justify expected exo benefits within ergonomics, or generate estimates that can be input into return-on-investment models.

A second alternative approach would map moment (or EMG) data onto Liberty Mutual Materials Handling (Snook) tables. This may be feasible; however, these tables reflect maximum acceptable weights due to physical exertion limits, and do not necessarily reflect injury risk to the back. To the extent that back strain is the main limiting factor in what intensity of lifting is acceptable, then this may also be a reasonable approach. But this is a critical assumption, and it remains unclear if or when it is well justified. Strain or fatigue of other muscles elsewhere in the body can also contribute to what workers subjectively perceive as acceptable levels of physical exertion. Additional investigation and validation of this approach would be needed, but progress could be challenging given the extensive experimental studies underlying the Liberty Mutual tables, the resources needed to repeat these types of experiments while wearing exos, and the issue of how to confirm generalizability of this approach for different types of back exos. Exo-LiFFT is better suited to assess injury risks because it unifies the etiology of back overexertion injuries and the biomechanical function of back exos.

A third alternative approach would be to adapt the revised NIOSH Lifting Equation, which uses various inputs (e.g., load constant, horizontal multiplier, distance multiplier, asymmetry multiplier) to compute a recommended weight limit and a lifting index, which provides a relative estimate of injury risk. This tool may also be feasible to adapt; however, a key challenge is that it is unclear how to modify this regression equation to accommodate exos. One option is to plug “equivalent weight” (when wearing an exo) into the revised NIOSH Lifting Equation. Or another option is to reduce the horizontal multiplier to account for the equivalent moment contribution from an exo. Both options predict a reduction in injury risk due to exo assistance, which is qualitatively consistent with our proposed Exo-LiFFT approach.

However, these two different modifications of the revised NIOSH Lifting Equation do not yield the same lifting index as each other, and it is not clear if one or the other option is more appropriate, or if other terms in the equation should also be updated due to exo assistance. Since the revised NIOSH Lifting Equation is a unitless regression, modification of this equation (or its inputs) is on ambiguous theoretical footing. This contrasts with the Exo-LiFFT equations, where the difference between the peak load moment and exo assistance moment has direct physical and physiological relevance, reflecting the reduction in biomechanical loading to a user’s back, and the associated reduction in tissue damage in accordance with the underlying etiology of back overexertion injuries. Additional investigation into modifying the NIOSH Lifting Equation for use with exos would be needed. New data with an exo may need to be collected, and a new regression equation fit to these data, potentially with additional assumptions and variables. Our assessment at this time is that the compatibility and validity of using the NIOSH Lifting Equation with exos, and justification for modifying the existing equations, remain less clear than using Exo-LiFFT.

#### A.2 Generalizability of Exo-LiFFT for different types of back exos

We expect Exo-LiFFT to be broadly applicable to different types of back exos (e.g., soft, rigid, passive, powered, Fig. 1) for the same reason existing ergonomic assessment tools have been generalizable to different material handling jobs and industries. Ergonomic assessment tools (e.g., revised NIOSH Lifting Equation, LiFFT) have been successful at distinguishing back injury risk using a crude (rough) estimate of loading on the low back; even without having access to tissue-specific or highly reliable estimates of lumbar tissue forces (Waters et al., 1993). For many ergonomic assessment tools compressive spine force is used as a general surrogate for back loading. However, it is not the compression of the spine that causes a back muscle to strain, ligament to tear, or necessarily even a disc to bulge (Sparto and Parnianpour, 1998). Spine compression force is most closely linked to vertebral endplate fractures and microfractures (Chaffin and Andersson, 1984; van Dieën et al., 1999; Waters et al., 1993). But since tensile forces in back extensors (e.g., muscles, ligaments) are the main source of spine compression force during bending and lifting (Lamers et al., 2018; McGill and Norman, 1986; Waters et al., 1993) these musculoskeletal loads all tend to increase (or decrease) together. For this reason, estimating spine compression force – or a correlated loading metric such as lumbar moment (McGill et al., 1996; Merryweather et al., 2009) – is useful in assessing or predicting LBD risks as a whole (Herrin et al., 1986; Waters et al., 1993).

Back exos of all types (soft, rigid, passive, powered; Fig. 1) exert a moment about the lumbar spine that offloads both tensile and compressive tissues of the user’s back (Kermavnar et al., 2020; Lamers and Zelik, 2021). The exo moment about the back can be thought of as an approximation of back offloading, which is used in the Exo-LiFFT equation to update the estimates of back loading on the user. In reality, whether one is wearing an exo or not, musculoskeletal dynamics (e.g., load sharing between tissues) is vastly more complex, and not currently measurable. Nevertheless, ergonomic assessment tools have shown empirically that even crude estimates of back loading -- when monitored across thousands of loading cycles per day, and hundreds of thousands of cycles per year -- can provide effective, useful and actionable insight on injury risk.

In the future, with sufficient advances in musculoskeletal modeling (e.g., that overcome challenges with reliably estimating musculoskeletal load distribution in the back) or non-invasive tissue-level load monitoring of musculoskeletal structures (Matijevich et al., 2021), it may be possible to provide separate risk assessments for back muscles vs. ligaments vs. vertebrae vs. spinal discs. But for now, simple ergonomics tools like LiFFT (and its exo-compatible version, Exo-LiFFT) provide contemporary options for assessing and predicting general back injury risk. As discussed herein, simulations can be used with Exo-LiFFT to compute a range of expected exo effects on back injury risk (e.g., from conservative to aggressive estimates). Ergonomic assessment tools can be further refined, validated, and calibrated in the future when more long-term epidemiological data or advanced monitoring tools become available.

